# Balancing revenue generation with capacity generation: Case distribution, financial impact and hospital capacity changes from cancelling or resuming elective surgeries in the US during COVID-19

**DOI:** 10.1101/2020.04.29.20066506

**Authors:** Joseph E. Tonna, Heidi A. Hanson, Jessica N. Cohan, Marta L. McCrum, Joshua J. Horns, Benjamin S. Brooke, Rupam Das, Brenna C. Kelly, Alexander John Campbell, James Hotaling

## Abstract

**Background:** To increase bed capacity and resources, hospitals have postponed elective surgeries, although the financial impact of this decision is unknown. We sought to report elective surgical case distribution, associated gross hospital earnings and regional hospital and intensive care unit (ICU) bed capacity as elective surgical cases are cancelled and then resumed under simulated trends of COVID-19 incidence.

**Methods:** A retrospective, cohort analysis was performed using insurance claims from 161 million enrollees from the MarketScan database from January 1, 2008 to December 31, 2017. COVID-19 cases were calculated using a generalized Richards model. Centers for Disease Control (CDC) reports on the number of hospitalized and intensive care patients by age were used to estimate the number of cases seen in the ICU, the reduction in elective surgeries and the financial impact of this from historic claims data, using a denominator of all inpatient revenue and outpatient surgeries.

**Results:** Assuming 5% infection prevalence, cancelling all elective procedures decreases ICU overcapacity from 340% to 270%, but these elective surgical cases contribute 78% (IQR 74, 80) (1.1 trillion (T) US dollars) to inpatient hospital plus outpatient surgical gross earnings per year. Musculoskeletal, circulatory and digestive category elective surgical cases compose 33% ($447B) of total revenue.

**Conclusions:** Procedures involving the musculoskeletal, cardiovascular and digestive system account for the largest loss of hospital gross earnings when elective surgery is postponed. As hospital bed capacity increases following the COVID-19 pandemic, restoring volume of these elective cases will help maintain revenue.

**DECLARATIONS:** *Ethics approval and consent to participate:* This study did not meet criteria for IRB review.

*Consent for publication:* Not applicable

*Availability of data and materials:* To facilitate research reproducibility, replicability, accuracy and transparency, the associated analytic code is available on the Open Science Foundation [1] (OSF) repository, [DOI 10.17605/OSF.IO/U53M4] at [https://osf.io/u53m4]. The data that support the findings of this study were obtained under license from Truven. Data were received de-identified in accordance with Section 164.514 of the Health Insurance Portability and Accountability Act (HIPAA).

*Competing interests:* JET received modest financial support for speakers fees from LivaNova and from Philips Healthcare, outside of the work. The other authors declare that they have no competing interests.

*Funding:* JET is supported by a career development award (K23HL141596) from the National Heart, Lung, And Blood Institute (NHLBI) of the National Institutes of Health (NIH). The content is solely the responsibility of the authors and does not necessarily represent the official views of the National Institutes of Health. None of the funding sources were involved in the design or conduct of the study, collection, management, analysis or interpretation of the data, or preparation, review or approval of the manuscript.

*Authors’ contributions:* JET, JH had full access to all the data in the study, takes responsibility for the integrity of the data, the accuracy of the data analysis, and the integrity of the submission as a whole, from inception to published article. JET, HH, BSB, JC, MM, JJH, JH conceived study design; JET, HH, BSB, JC, MM, JJH, RD, BK, AJC, JH contributed to data acquisition and analysis; JET, HH, JJH, JH drafted the work; all authors revised the article for important intellectual content, had final approval of the work to be published, and agree to be accountable to for all aspects of the work.

*Acknowledgements:* Not applicable

## INTRODUCTION

The novel Coronavirus Infectious Disease 2019 (COVID-19) is a highly transmittable virus that has resulted in over 2.5 million infections worldwide [2]. United States (US) health systems are designed for stable, predictable utilization patterns; they are generally unprepared for surge medical need as is required with the COVID-19 pandemic. Surge situations have historically been addressed by deploying temporary medical teams and tent facilities, or have overwhelmed the available resources with resultant excess morbidity and mortality [3]. Current estimates suggest 5% of the US population will contract COVID-19, of whom 15% will require hospitalization, and 5% will require intensive care [4–6]. At this rate, there will be 5 infected patients requiring hospitalization for every existing US hospital bed in addition to concomitant hospitalized non-COVID patients [7].

In order to decrease the resources needed during the peak of COVID-19, many countries have adopted physical distancing to attempt to flatten the curve of COVID-19 [8]. Some nations, such as China, have taken extreme measures such as forced quarantines and a completely shutdown of all aspects of society whereas other countries such as Norway have chosen to remain largely open. The United States has emphasized physical distancing and suspension of non-essential business operations which have, until now, included elective surgical procedures. This has resulted in hospitals in areas with relatively few COVID-19 patients, such as Utah, functioning at only 30% of their typical inpatient and surgical capacity. The financial implications of cancelling elective surgical procedures has been devastating for healthcare systems with some systems currently losing upwards of $25 million USD per week [9].

However, to mitigate large anticipated surges in hospitalizations and deaths over the coming months, US hospitals need to sustainably absorb increases in patients requiring hospitalization and intensive care, while continuing to care for non-COVID patients. After a statement from the American College of Surgeons (ACS), many hospitals across the country ceased elective surgery to free beds and limit patient exposure. While cancelling elective surgical cases has increased capacity, the financial impact of these surgeries, including relative financial contribution by case type, has not been described. Furthermore, the epidemiology of the deferred elective surgical cases, case counts, and their resultant contribution to bed capacity has not been described.

We estimate the financial implications of cancelling elective surgeries using a national insurance claim database, Truven MarketScan (MS). This information can be used to inform strategies for resuming elective surgical procedures. We describe regional differences in capacity of US hospitals to absorb COVID-related inpatient surges if measures are not taken to slow the spread, describe elective case distribution, and present the relative financial impact and case counts of resumption of elective surgical cases.

## METHODS

Our analysis is reported according to the Strengthening the Reporting of Observational Studies in Epidemiology (STROBE) Guidelines (Supplemental Methods) [10].

### Data Source and Study Population

Gross inpatient hospital earnings, outpatient surgical earnings, and inpatient and ICU (I/ICU) beds were estimated using Truven MarketScan (MS). MS billing data captured 8-16% of the entire US population from 2013–2017. For every MS patient, we determined the total number of I/ICU days from 2008–2017 for 161 million (M) enrollees and aggregated visit counts by month, state, and major diagnostic category (MDC). Elective surgery admissions were identified by surgery revenue codes not associated with emergency revenue or provider codes. Admissions occurring within 30 days post-elective (including outpatient) surgery were considered complications and categorized as resulting from elective surgery. Gross earnings were aggregated by MDC codes and surgical (elective vs non-elective) vs non-surgical. Analysis by MDC excluded eye, human immunodeficiency virus, health status and missing categories.

### Study Variables and Outcomes

MS data were used to report gross hospital earnings derived from inpatient admissions and outpatient surgeries for each MDC category by elective vs non-elective surgical and non-surgical. We calculated proportion of gross earnings from elective surgeries by state, and the proportion of I/ICU beds unoccupied, and occupied by elective and emergent cases by state. The average number of unoccupied beds was defined as the difference between the average number of I/ICU days and the maximum number of I/ICU days in a given state. We used data from the Harvard Global Health Institute (HGHI) to identify the total number of I/ICU patient beds per state and applied the proportion from the MS data to obtain estimates of occupied beds [11].

The Centers for Disease Control (CDC) Morbidity and Mortality Weekly Report (MMWR) was used to calculate the number of expected I/ICU beds occupied by age group [12]. We applied these age-specific rates to 5% of each state’s population using 2018 U.S. Census Bureau age-specific population estimates to calculate the number of expected hospitalizations and inpatient cases by state [13]. We calculated lower bound (LB) and upper bound (UB) estimates based on the CDC’s reported uncertainty.

### Statistical Analysis

In order to estimate the financial implications of cancelling or resuming elective surgical cases, we calculated the distribution of gross earnings by MDC code and then by elective and non-elective surgical, and non-surgical classifications. To inform the balance of revenue and capacity generation, we illustrate the effect of cancellation of elective surgical cases on hospital capacity by modeling a scenario that may exist if 5% of the US population were infected with COVID-19. We constructed a Generalized Richards Growth Model (GRM), which has been proven effective for capturing single epidemic transmission dynamics, to describe a scenario that could happen in the absence of limitation to the disease spread [14]. Our GRM assumed a 5% infection rate, 5.2-day doubling rate, and 100-day period of disease progression. These values were based on current published rates and do not account for variation in social distancing [15, 16].

We illustrated gross earnings by MDC code and by classification. We calculated the number of hospitalized patients in each state during the simulated pandemic peak using 11-day inpatient and 9-day ICU length of stay [5, 6, 17]. We then calculated the ratio of cases to total number of available beds per state with and without removal of elective surgery.

## RESULTS

Across the U.S. there were an average of 1,442,013 (Interquartile range [IQR]: 1,378,039, 1,507,994) inpatient days per month and 104,265 ([IQR] 101,961, 104,842) ICU days per month in the MS sample. Roughly 30% of these days were associated with elective surgery (**Figure 1**). We applied these percentages to US Hospital data and classified beds as emergent, elective, and available. Of 735,996 hospital beds in the US, 351,369 were emergent, 136,264 were elective, and 248,363 were available.

**Figure 1.**
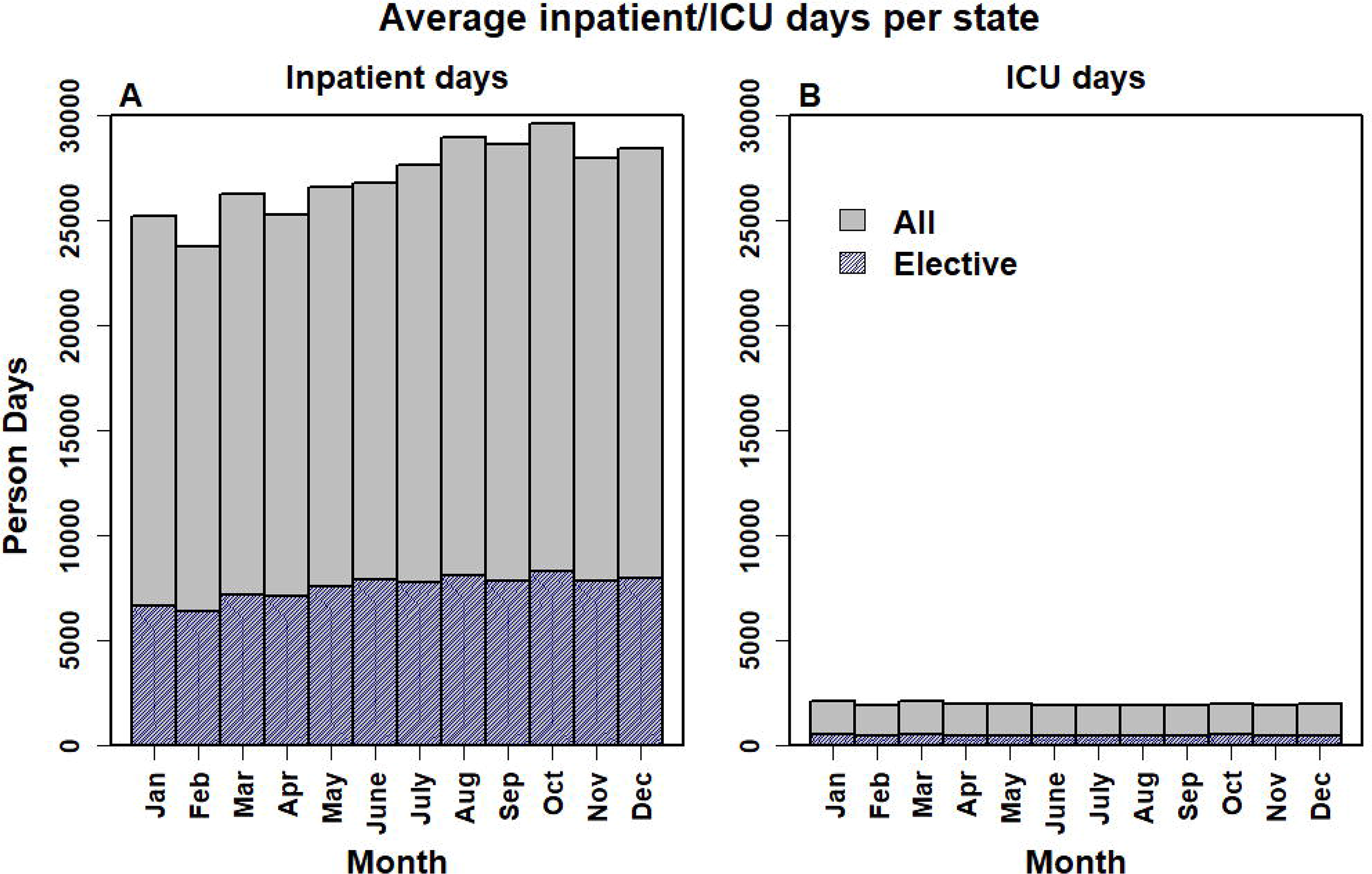
Average Proportion of Inpatient and ICU Person Days per State Resulting from Elective Surgery. Data from 161 million Marketscan patients from 2008–2017 displaying aggregated counts of hospital and ICU beds in total and those resulting from elective surgery averaged across all states. For each month.

### Financial Impact

Elective surgical cases contribute 78% (IQR of statewide variation 74, 80) to of the gross inpatient and outpatient surgical earnings per year for hospitals represented in our sample, or 1.1 trillion US dollars. This value varies by MDC from $488M to $231 billion (B) (median 39B; IQR 16B, 59B) (**Figure 2**). Within each MDC, the percent of earnings from elective surgical cases varies from <27% to 97% (median 68%; IQR 53, 86). When restricting our analyses to inpatient-derived revenue, elective surgery accounted for 43% (IQR of statewide variation 40, 45) of total gross earnings, or $254B. This value varies by MDC from <1% to 88%, ($25M to $80B). The relative financial contribution of elective surgical cases varies regionally and has important financial implications for hospital specific decisions on resumption of these elective procedures (**Figure 3**).

**Figure 2.**
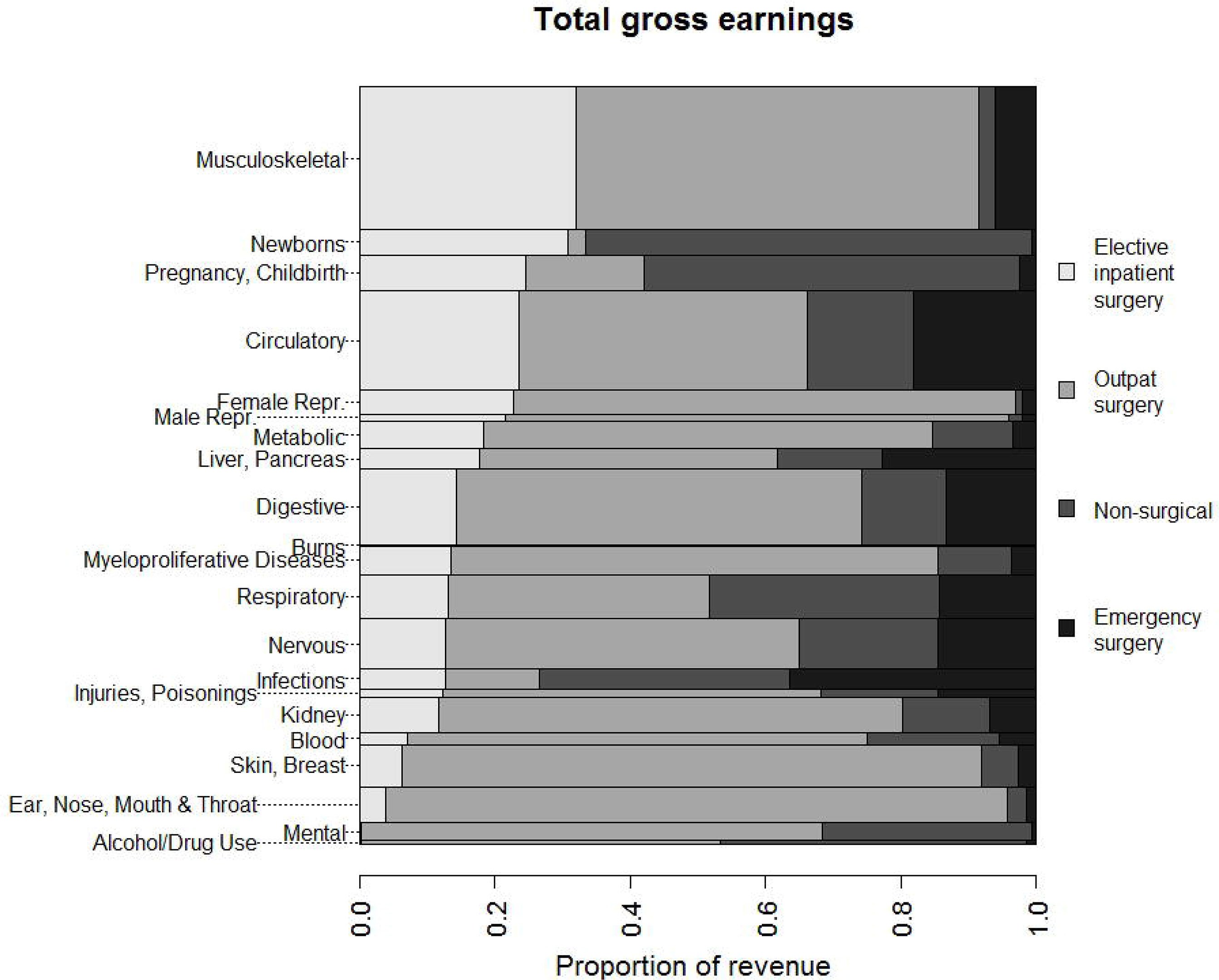
Financial contribution of major diagnostic categories (MDC) to gross hospital earnings. Data from 161 million Marketscan patients from 2008–2017 displaying aggregated gross hospital earnings by surgery type, separated by major diagnostic category (MDC), across the US. Levels are listed in descending order the percentage of each MDC category contributed by elective inpatient cases. Level width is proportional to the absolute value in US dollars.

**Figure 3.**
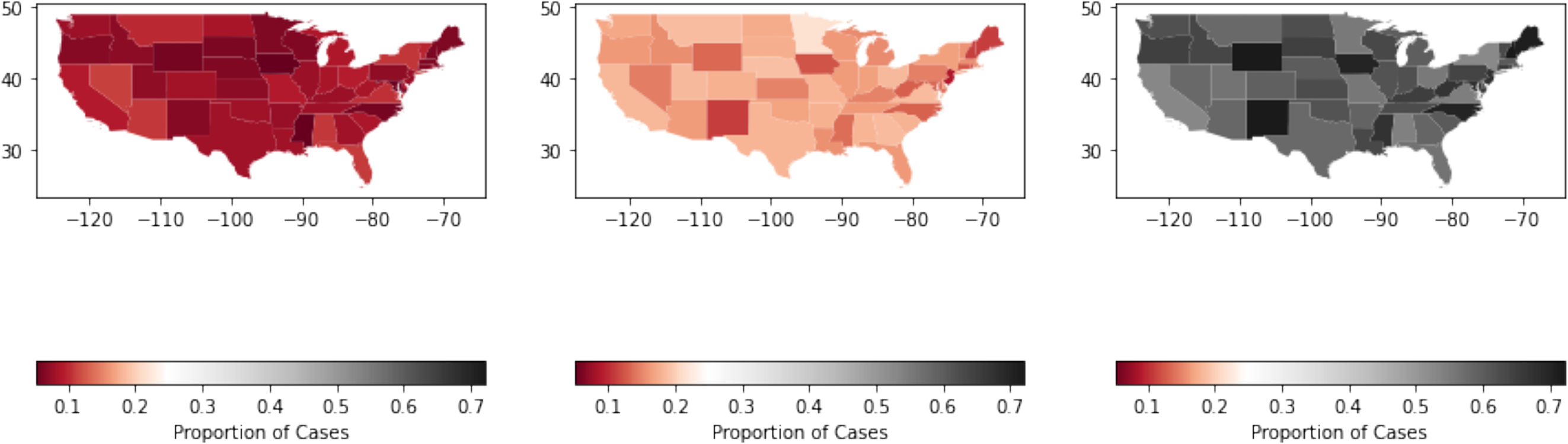
Regional variation by state in percentage financial contribution of non-elective, elective inpatient and outpatient surgeries. **Panel A** shows percent financial contribution to gross hospital earnings by state for non-elective cases. **Panel B** shows elective inpatient cases. **Panel C** shows outpatient cases.

### Case Counts

With the cancellation of elective surgery, there is a need to balance revenue generation with capacity increases. Elective surgical procedures contribute 3B cases in the MS population, of which 7.9M are inpatient. This varies by MDC (median 96M [IQR 47M, 224M], range 1.9M – 515M), of which 197396 (IQR 86781, 372544; range 915 - 2.4M) are inpatient. Elective musculoskeletal, circulatory and digestive categories comprise 1.1B cases, of which 3.8M are inpatient. These three categories together compose 13% of hospital admissions, but 23% ($142B) of inpatient gross earnings. Including outpatient surgical cases, they compose 33% ($447B) of total inpatient and surgical-outpatient hospital earnings.

### Hospital and ICU Capacity

Cancelling elective surgeries adds significant capacity for *new daily ICU admits* (**Figure 4A**), but falls short of the *full capacity required* when 5% of the population is infected over a relatively short period of time and median ICU length of stay is 9 days (**Figure 4B**). Without cancelling elective surgery, median state hospital and ICU capacity was 4967 (IQR 1867, 6100) beds, and 743 (IQR 254.94, 944.37) respectively. This increased to 7692 (IQR 2553, 9679) hospital beds, and 991 (IQR 298, 1197) ICU beds if elective surgeries were cancelled.

**Figure 4.**
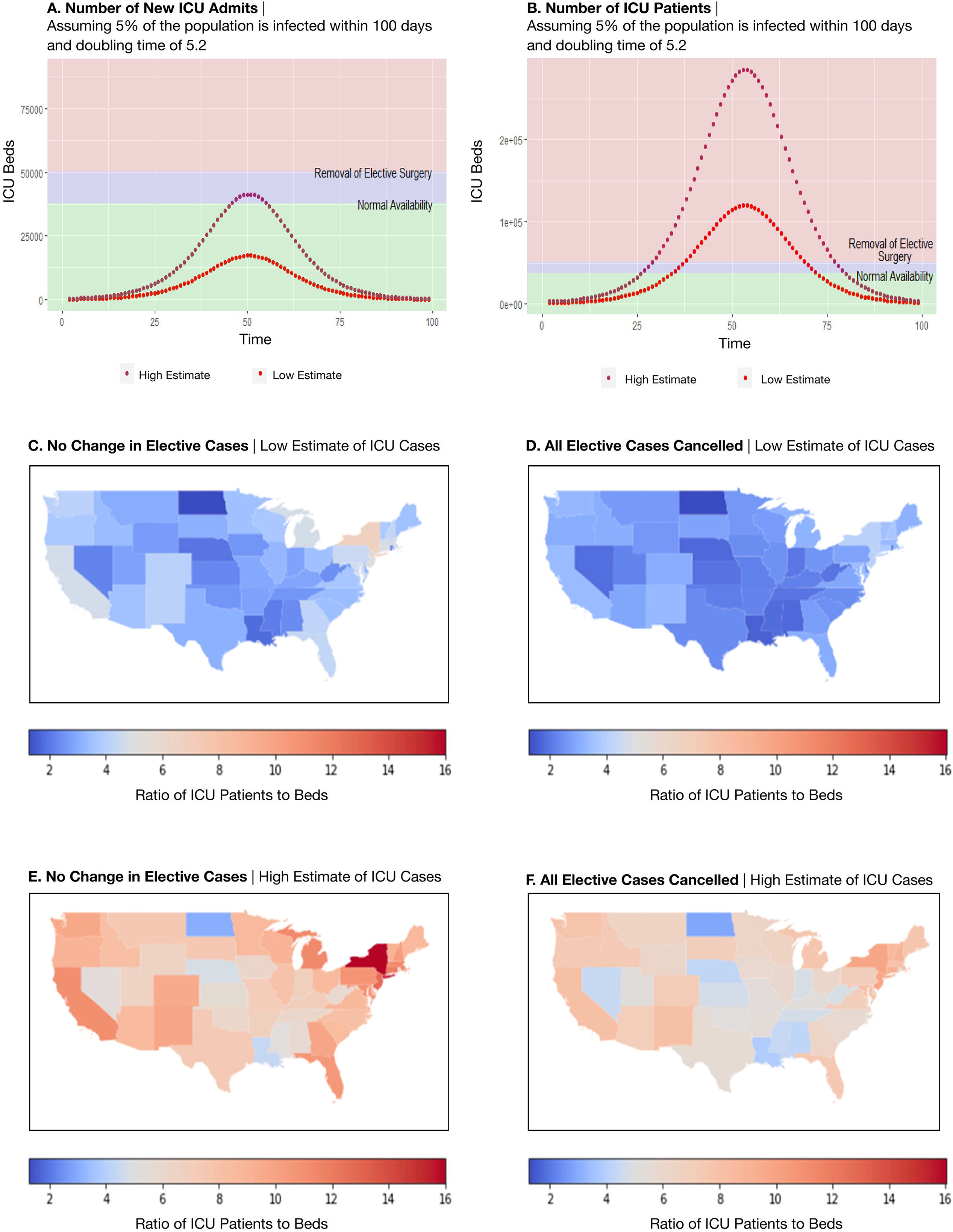
ICU Capacity across the US with and without cancelling elective surgeries. **Panel A** represents the number of ICU patients admitted each day while **Panel B** represents total ICU patient days during peak infection assuming each COVID-19 patient spent 9 days in the ICU. Green denotes normal availability of ICU beds, purple shows the additive ICU beds that are available when all elective surgeries are cancelled and pink indicates ICU capacity that would have to be generated by adding additional capacity in hospitals beyond what could be generated through cancellation of elective cases. The dotted lines represent estimates of the number of patients admitted into ICUs from the time the first person was infected and modeled for 100 days after this based on a Generalized Richards Growth Model (GRM). The low and high estimates for COVID-19 infection are based on the CDC low and high estimates. **Panels C-F:** Impact of Cancelling All Elective OR Cases on ICU Bed Availability if 5% of U.S. Population Infected with COVID-19. Estimates of low (**Panel C**) and high (**Panel E**) ICU cases are derived the uncertainty estimates from CDC MMWR reports from the CDC COVID-19 Response Team. Additional capacity through cancellation of elective cases (**Panel D, Panel F**) was determined by applying estimates of the occupied and unoccupied beds resulting from elective surgery from the Marketscan database to the Harvard Global Health Institute (HGHI) estimates of total inpatient and ICU beds in each state.

There is variability in hospital capacity by state. As modeled, states would have mean 3.4 patients requiring admission per available ICU bed (IQR: 2.65 – 4.09) (**Figure 4C**). If we use the UB rates of ICU provided by the CDC, this increases to mean 8.2 patients per bed (IQR: 6.33 – 9.67) (**Figure 4E**). Removing elective surgeries will alleviate some of this stress, with LB estimates of an average of 2.7 ICU patients per bed IQR (2.19 – 3.16) (**Figure 4D**) and UB estimates of an average 6.4 (IQR 5.3 – 7.6) (**Figure 4F**).

## DISCUSSION

Using billing and utilization data to model the financial contribution of elective surgical cases to hospital gross earnings, we demonstrate that cessation of elective musculoskeletal, cardiovascular and digestive cases account for the largest loss of hospital gross earnings, at $447B or 33% of all inpatient and surgical-outpatient revenue. In contrast, by case count, musculoskeletal, pregnancy, and circulatory categories account for the greatest contribution of elective inpatient cases. Applying case counts to the available hospital beds, cancelling elective surgeries at all US hospitals will decrease ICU overcapacity from 340% to 270% (lower bound) or from 820% to 640% (upper bound) assuming 5% of the U.S. population is infected as shown in our models, but at a financial cost. In light of the significant contribution of elective surgeries to gross hospital earnings, selective resumption of high contributing elective MDC categories may be a way to resume surgery in a financially sustainable way when deemed safe.

Continued provision of care to patients with COVID-19 involves balancing increasing healthcare capacity with sustaining revenue generation. Previous models have addressed the expected healthcare burden of COVID-19 in various ways, but have not addressed revenue generation. To our knowledge, ours is the first to report case distribution and financial contribution of elective surgeries. We incorporated state specific billing data, reporting the relative contribution of elective surgeries to gross hospital earnings, and case counts, by MDC code. Our study informs the relative financial and capacity impact from selective resumption of elective cases. Further, we model the expected increase in bed capacity from cancelling these surgeries. Tsai *et al* previously estimated 1.7 excess COVID-19 patients per hospital bed and 4 excess patients per critical care bed assuming a 40% cumulative infection rate over 12 and 18 months, and an aggressive 50% bed availability [18]. Similarly, Murry used occupancy data from Medicare and Medicaid patients to estimate a capacity gap of 17,000 ICU beds and 64,000 hospital beds at peak infection assuming some measure of social distancing [19]. Our study differs from these by modeling disease transmission under a lower infection rate (5%) but faster course (100 days) than Tsai *et al*, and in a worse-case scenario without complete shelter in place orders, which 9 states do not currently have [20]. We also incorporated state-specific data for elective case volume and temporal variation. We demonstrated substantial state-level variation in overcapacity. This suggests an opportunity for regional cooperation and resource redistribution.

Our results are generated using a financial denominator of total inpatient revenue plus outpatient surgical cases, and do not account for the Medicare population that is not insured by Part C. We determined overcapacity values using consistent rates of infection across the population in less than 100 days and assumptions that regional rates are equal [7, 18, 19]. It is important to note that we applied rates of disease progression from regions prior to enforced distancing. Consistent with this, we assumed a stable rate of disease progression of 100 days since first case and do not account for policy responses to the pandemic, such as closure of non-essential service and stay-at-home orders, which are highly variable across states.

## Conclusions

Elective inpatient surgeries account for and account 27% of hospital and ICU beds and 43% of gross earnings, which varies substantially by specialty. Among elective cases, musculoskeletal, cardiovascular and digestive MDC categories account for the largest contribution to hospital gross earnings, at 33%. The greatest contribution of bed capacity comes from musculoskeletal, pregnancy and circulatory categories. The cancellation of elective surgery will result in a substantial increase in hospital and ICU bed capacity, though this will vary between states, and at significant financial cost.

## Data Availability

To facilitate research reproducibility, replicability, accuracy and transparency, the associated analytic code will be made available upon publication after peer review on the Open Science Foundation [1] (OSF) repository, [DOI 10.17605/OSF.IO/U53M4] at [https://osf.io/u53m4]. The data that support the findings of this study were obtained under license from Truven. Data were received de-identified in accordance with Section 164.514 of the Health Insurance Portability and Accountability Act (HIPAA).

